# Should Multi-Cancer Early Detection Testing Replace Guideline-Recommended Colorectal Cancer Screening? A Comparative Modeling Analysis

**DOI:** 10.64898/2026.07.10.26357782

**Authors:** Ishfaq Ahmad, Carolyn M. Rutter, Chris E. Maerzluft, Isabel Dengos, Kemal C. Gogebakan, Jane M. Lange

## Abstract

**Background:** Colorectal cancer (CRC) screening strategies such as colonoscopy and fecal immunochemical testing (FIT) reduce CRC mortality through both early detection and prevention via precursor lesion removal. Multicancer early detection (MCED) blood tests offer the potential to detect multiple cancers with a single assay but provide little opportunity for cancer prevention. Whether the ability to detect multiple cancers can offset the loss of CRC prevention remains unclear.

**Methods:** We used microsimulation to compare MCED and guideline-recommended CRC screening strategies. CRC outcomes were simulated using CRC-SPIN v3.0 and non-CRC cancers using MCEDsim, calibrated to SEER incidence data. Assuming optimistic MCED preclinical sensitivity equal to published case-control estimates, we compared life-years gained and late-stage disease outcomes for annual FIT, decennial colonoscopy, and MCED-only strategies across a range of preclinical durations and survival benefit assumptions.

**Results:** Relative to no screening, colonoscopy and FIT reduced late-stage diagnoses by 26% and 25%, respectively, versus 20%–32% for annual MCED screening. Across assumptions, MCED-only strategies generated 33%–51% as many life-years gained as colonoscopy.

**Conclusions:** Currently available MCED tests are unlikely to be effective replacements for guideline-recommended CRC screening, which derives substantial benefit from the detection and removal of precursor lesions. MCED screening may provide additional benefit as a supplement to recommended CRC screening.

## 1 Introduction

Despite substantial progress in prevention and early detection, colorectal cancer (CRC) remains a leading cause of cancer-related morbidity and mortality in the United States.^1^ Current CRC screening guidelines from the U.S. Preventive Services Task Force (USPSTF)^2^ and the U.S. Multi-Society Task Force on Colorectal Cancer^3^ recommend a number of evidence-based screening strategies, including decennial colonoscopy and annual fecal immunochemical testing (FIT) with follow-up colonoscopy for positive results. These approaches have demonstrated substantial preventive benefit by detecting CRC at earlier stages and by identifying and removing precancerous adenomas and sessile serrated lesions (SSLs). While FIT and colonoscopy have limitations and harms,^4, 5, 6, 7^ empirical^8, 9^ and modeling studies^10^ have demonstrated that guideline-recommended screening strategies substantially reduce CRC mortality and life-years lost from colorectal cancer.

Recent advances in biomarker technologies have driven the development and market introduction of blood tests for CRC, with the goal of increasing screening uptake among currently unscreened individuals.^10^ However, unlike guideline-recommended strategies, these tests provide little opportunity for cancer prevention because they detect precursor lesions only rarely; published data on methylation-based blood tests show advanced adenoma sensitivities of 11–13%, roughly equivalent to their false-positive rates of approximately 10%.^11, 12^ Thus, these tests offer limited preventive benefit and lower effectiveness than FIT or colonoscopy; modeling studies suggest that screening based on CRC blood tests yields only 85–87% of the life-years gained from colonoscopy^13^ while cost-effectiveness analyses indicate that currently reimbursable tests save fewer lives at higher cost than guideline-recommended strategies.^14^

In parallel, multicancer early detection (MCED) blood tests have emerged as a novel screening paradigm, leveraging circulating tumor-derived signals similar to those used in CRC blood tests, including DNA methylation patterns.^15^ Existing MCED assays aim to detect multiple solid tumors using a minimally invasive blood test and are designed to achieve very high specificity.^16, 17^ Because adenoma detection by methylation-based blood tests appears to occur largely through nonspecific test positivity, the high specificity of current MCED assays suggests that adenoma detection rates are likely negligible.

As MCED tests enter clinical practice, it is unclear how their benefits and harms compare with those of established screening programs supported by substantial evidence of efficacy. For CRC, a key unresolved question is whether replacing guideline-recommended CRC screening with MCED testing would result in a net gain or net loss in lives saved at the population level. Specifically, does the additional benefit of detecting multiple cancers, beyond just CRC, outweigh the loss of benefit associated with no longer preventing CRC through precursor lesion removal?

To address this question, we use a microsimulation modeling approach that combines the validated Colorectal Cancer Simulated Population model for Incidence and Natural History (CRC-SPIN v3.0)^18^ with a newly developed multicancer early detection simulation framework, MCEDsim, based on previously published analytic models of multicancer detection testing.^19^ The CRC-SPIN model has been extensively used to inform policy decisions, including contributing to the evidence base underlying U.S. Preventive Services Task Force recommendations for colorectal cancer screening.^10, 2^ The combined modeling framework allows us to quantify and compare key features of screening benefit across MCED-based and guideline-recommended CRC screening strategies, including life-years gained and reductions in late-stage disease at diagnosis.

### 1.1 Overview

The microsimulation study used a parallel universe approach to examine the benefits of counterfactual CRC and MCED screening strategies across a common set of individuals. We assumed an MCED test that detects the following solid cancer sites: anus, bladder, colorectal, esophagus, gastric, head and neck, liver, lung, lymphoma, ovary, pancreas, renal, and uterine. These cancers include the solid tumor types evaluated in the NHS-Galleri validation study^20^ and the eight cancer types targeted by CancerSeek^21^ (currently CancerGuard^TM^). Breast and prostate cancers were not included due to guideline-recommended screening for breast cancer and poor sensitivity of existing MCED tests for prostate cancer.^15^

We simulated a cohort of 10 million individuals (5 million per sex), with cancer risk that is representative of the general U.S. population. Individuals entered the simulation at age 45 (calendar year 2026) and underwent screening from age 45 to 75. For each individual, natural histories for all targeted cancer sites were simulated independently, and outcomes were simulated until death, with a maximum age of 100 years. Other-cause mortality was simulated using inverse-transform sampling based on adjusted all-cause mortality life tables that excluded deaths attributable to CRC and MCED-targeted cancers. Each individual was allowed to develop at most one cancer, either CRC or one of the MCED-targeted cancers, over their lifetime. To ensure that simulated incidence matched marginal population rates, subsequent primary cancers were allocated to individuals who remained free of clinically detected cancer over their lifetime, matched on age at other-cause death. We evaluated 6 screening strategies (Table 1).

**Table 1:**
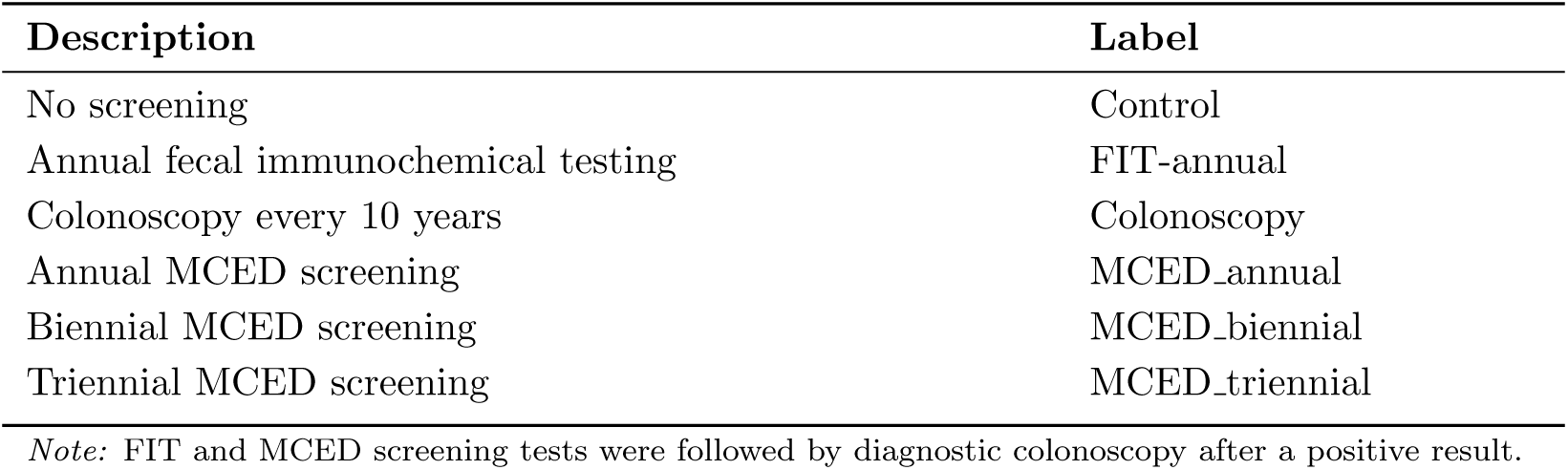
Description of the 6 population screening strategies evaluated.

#### 1.1.1 Outcomes

Screening strategies were evaluated based on total life years and the cumulative incidence of 1) total cancer diagnosed within a person’s lifetime 2) cancer diagnosed in late stage, where late stage generally corresponds to AJCC-7 stage III-IV.^22^ Life-years were defined as the minimum of age at other-cause death and age at cancer-specific death. Life-years gained (LYG) were computed based on differences between control and screening strategies. Harms were quantified using the rate of false-positive screening examinations for MCED strategies. We did not explicitly track false-positive burden for FIT-based strategies because downstream colonoscopy following a positive FIT serves as both diagnostic confirmation and an opportunity for lesion removal.

### 1.2 Simulation models

#### 1.2.1 CRC-SPIN

The Colorectal Cancer Simulated Population model for Incidence and Natural history (CRC-SPIN v3.0) simulates development of colorectal cancer (CRC) in adults, arising through both adenomas and SSLs^23^ (Supplemental Figure 1). CRC-SPIN includes birth-cohort effects that capture recent increases in CRC incidence.^24, 25, 26^ The model simulates events in continuous time, including onset of adenomas and SSLs via a Poisson process, lesion growth, and transition to preclinical CRC based on lesion size, time from transition to preclinical cancer to clinical detection, stage at clinical detection, stage at screen-detection (when this occurs), and death from CRC and other causes. For this application, the model simulated stage-specific CRC survival and other cause survival that excluded death from other cancers simulated using the MCEDsim model (described in section 1.3 below). Individuals may develop multiple preclinical lesions, each with the potential to develop into CRC, though most do not make this transition. Individual-level outcomes are driven by the first CRC detected (clinically or via screening). CRC-SPIN v3.0 was calibrated to multiple targets that describe precursor lesion prevalence, size and multiplicity by age and sex, and CRC incidence, and externally validated to screening and surveillance studies.^23^

Simulated screening can detect and remove precursor lesions, preventing CRC, and can detect preclinical cancer leading to potentially earlier stage at detection. However, removal of precursor lesions may prevent one CRC, but allow another to arise.

#### 1.2.2 MCEDsim

Natural histories for non-CRC MCED-targeted cancers were generated using MCEDsim, which relies on a five-state natural history framework (Supplemental Figure 2) calibrated separately for each solid cancer site included in the MCED test.^19^ In this framework, individuals begin in a healthy state at age 0, progress to a preclinical detectable state, and ultimately to symptomatic, clinically detected disease. The preclinical state is subdivided into early and late stages. In general, early corresponds to AJCC-7^22^ stages I–II and late to stages III–IV; for pancreatic cancer, early is defined as stage I and late as stages II–IV, reflecting resectability considerations.

Dwell times in the early and late preclinical states are assumed to follow exponential distributions, with constant transition rates between preclinical and clinical states. The age at preclinical onset is parameterized using a sequence of exponential transitions that allow for a flexible hazard function increasing with age.

For each individual, given a simulated natural history of primary cancer, screening was superimposed at specified intervals using site- and stage-specific sensitivities (Table 3) to determine age and stage at screen detection. MCEDsim assumes that MCED-targeted cancer–specific survival is driven by stage at detection: cancers detected at an earlier stage through screening receive an increase in cancer-specific life expectancy corresponding to the difference in survival between early- and late-stage diagnoses in the absence of screening. This stage-shift assumption is commonly used in mechanistic models of cancer screening benefit.^27^ Operationally, post–lead-time survival late as simulated for each individual based on age, sex, and stage at diagnosis (early versus late) using inverse-transform sampling. In this way, individuals diagnosed at an early stage under screening have survival projected assuming early stage at the point of clinical diagnosis, regardless of their stage at diagnosis in the absence of screening.

#### 1.2.3 Calibration of MCEDsim

MCEDsim natural history models were calibrated using sex-, site-, age-, and stage-specific incidence data from the SEER Research Data (17 registries, years 2010–2021),^28^ following the methods described in Lange et al.^19^ Cancer sites were identified using the SEER Site and Morphology variable based on the ICD-O-3 Site Recode (2023 revision).

AJCC 7th edition staging information is unavailable in SEER data after 2015. For calendar years 2016–2021, we therefore imputed stage-specific incidence rates for each sex, site, and age group by combining overall incidence with sex-, site-, and age-specific proportions of early- and late-stage cases estimated from the 2010–2015 period.

Incidence data alone are insufficient to identify all transition rates between preclinical and clinical disease states. Indeed, the natural history models require specification of two key parameters: the overall detectable mean sojourn time (OMST), defined as the average duration of detectable preclinical disease, and the late-stage detectable mean sojourn time (LMST), defined as the average duration of detectable late-stage disease prior to clinical diagnosis. Varying these parameters allows the models to accommodate different assumptions regarding the length of the preclinical detectable window. The specific choices for OMST and LMST are discussed in Section 1.5.

### 1.3 Cause-specific survival models

Cause-specific survival times for CRC and MCED-targeted cancers were simulated using log-logistic survival models fit to case-listing cause-specific survival data from the SEER Research Data (17 registries, years 2010–2015). Cancer sites were identified using the SEER Site and Morphology variable based on the ICD-O-3/WHO 2008 Site Recode. Models were fit stratified by sex, and AJCC 7th edition stage (I–IV for CRC and early versus late stage for all other sites, as previously defined).

### 1.4 Other-cause death

Other-cause death was defined as death due to any cause other than CRC or the MCED-targeted cancers and was simulated based on custom other-cause mortality life tables that remove the risk of death from

CRC and MCED-targeted cancers. To construct these life tables, we used data from the CDC WONDER database (“2018–2022: Underlying Cause of Death by Single Race Categories”), which provides total deaths and population counts by age, sex, calendar year, and cause of death. Because population counts are not reported for ages 85 and older in CDC WONDER, we used life tables from the Human Mortality Database to impute population counts for ages 85–100 for calendar years 2018–2023.

To exclude CRC and MCED-targeted cancer deaths from all-cause mortality life tables, we applied the actuarial table adjustment approach described by Rosenberg.^29^ Supplemental Figure 3 compares the resulting other-cause mortality rates with all-cause mortality rates across age groups and by sex, illustrating the impact of excluding MCED-targeted cancer deaths from the overall mortality profile.

### 1.5 Test performance assumptions

Tables 2 and 3 present the sensitivity inputs used for FIT, colonoscopy, and MCED screening. FIT sensitivity is at an individual-level and is based on cancer sensitivity if preclinical cancer is present at the time of screening; otherwise sensitivity is based on the size of the largest adenoma present. Colonoscopy sensitivity is at the lesion-level and is based on lesion type and, for adenomas and SSLs, lesion size. MCED detection of CRC is based on the most advanced preclinical cancer with different sensitivity for early (Stage I,II) vs late (Stage III, IV) CRC. We assume that MCED tests have no ability to detect CRC precursor lesions, including adenomas and sessile serrated lesions, though positive MCED results are possible in people with precursor lesions but not CRC based on the test’s false positive rate. For non-CRC cancer sites targeted by the MCED test, sensitivity is specified for early and late stage.

**Table 2:** Test sensitivity inputs for CRC-related outcomes by screening modality. MCED sensitivity for CRC is specified separately for early- and late-stage disease based on case–control estimates from the CCGA study.

| **Outcome** | **FIT** | **Colonoscopy** | **MCED** |
| --- | --- | --- | --- |
| Adenomas <10 mm | 0.08 | 0.61–0.80 | 0.00 |
| Adenomas ≥10 mm | 0.24 | 0.91 | 0.00 |
| Sessile serrated lesions (SSLs) | 0.00 | 0.10–0.50 | 0.00 |
| CRC | 0.74 | 0.91 | Early: 0.67; Late: 0.92 |

MCED sensitivity inputs for both CRC and non-CRC cancer sites on the GRAIL’s Galleri test as an exemplar. The Circulating Cell-free Genome Atlas (CCGA) case-control study for GRAIL’s Galleri test^15^ estimated sensitivity in clinically-diagnosed individuals. To represent an optimistic assumption regarding screening efficacy, early and late-stage preclinical sensitivity is set equal to the CCGA estimates (Table 3.)

**Table 3:**
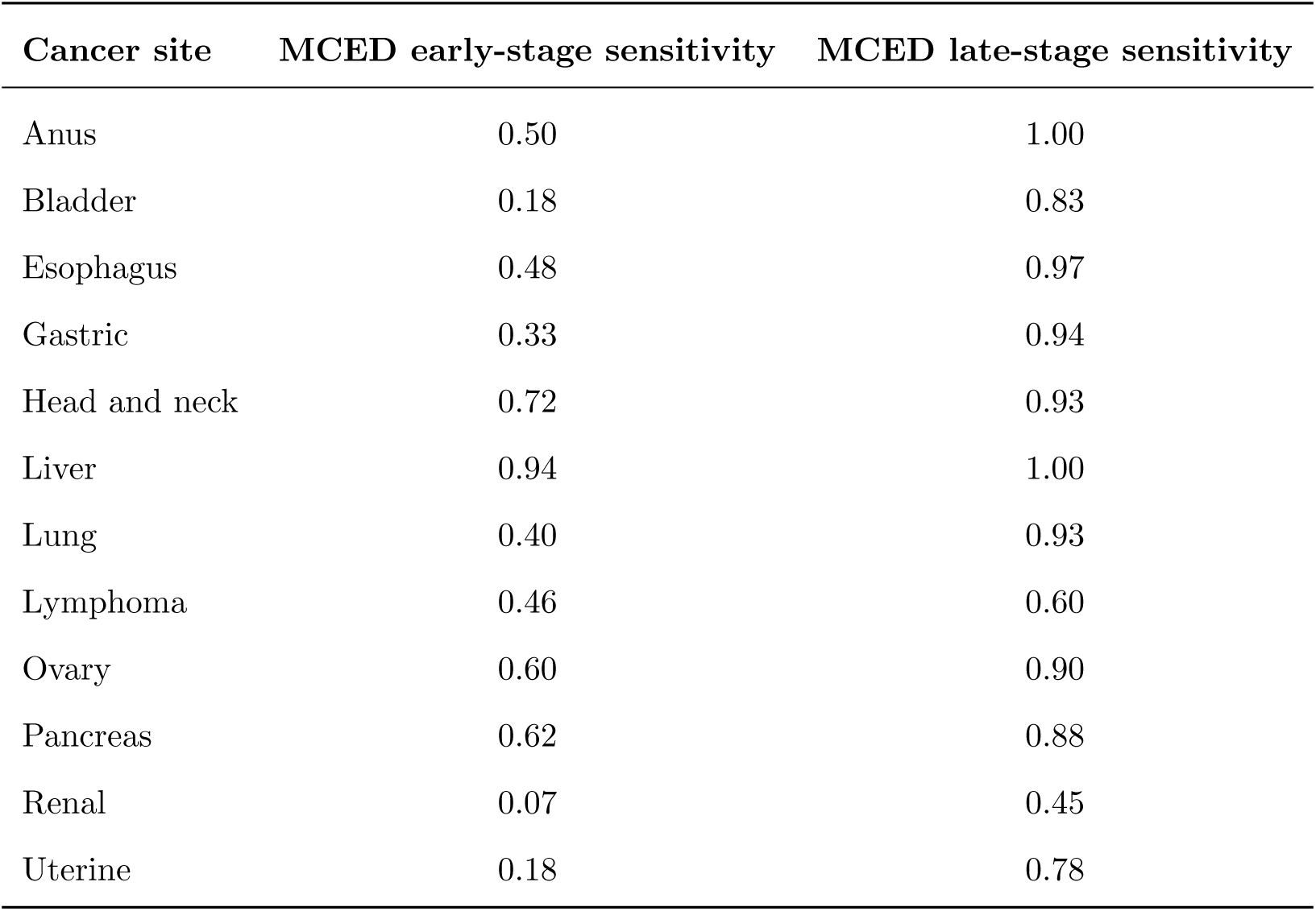
Stage-specific MCED test sensitivity inputs for non-CRC cancers, based on Klein et al. ^15^

| **Cancer site** | **MCED early-stage sensitivity** | **MCED late-stage sensitivity** |
| --- | --- | --- |
| Anus | 0.50 | 1.00 |
| Bladder | 0.18 | 0.83 |
| Esophagus | 0.48 | 0.97 |
| Gastric | 0.33 | 0.94 |
| Head and neck | 0.72 | 0.93 |
| Liver | 0.94 | 1.00 |
| Lung | 0.40 | 0.93 |
| Lymphoma | 0.46 | 0.60 |
| Ovary | 0.60 | 0.90 |
| Pancreas | 0.62 | 0.88 |
| Renal | 0.07 | 0.45 |
| Uterine | 0.18 | 0.78 |

#### 1.5.1 False positive assumptions

FIT tests may results in a false positive finding when patients without cancer or a preclinical lesion are sent to follow-up colonoscopy. These patients are assumed to return to FIT testing in 10 years. For these analyses, we do not consider false positive colonoscopy results, which occur when patients undergo biopsy that is later found to be benign tissue (a clean colonsocopy, i.e., neither a precursor lesion nor a preclincal CRC). False positive results from colonoscopy primarily affect costs, which we do not consider.

For MCED screening, we assume a per-screen false positive rate of 0.5%, consistent with the Galleri test’s specificity reported in the CCGA validation study.^15^ If the MCED test indicates CRC, then the patient undergoes colonoscopy. Even if the colonoscopy is “clean,” these patients return to MCED screening because the intent is to screen for a broader range of cancers. For each individual, the number of false positive results over the screening period was modeled as a binomial random variable with parameters *n* (the number of screening tests completed prior to cancer onset) and *p* = 0.005 (the per-screen false positive probability). Thus, individuals may experience multiple false positive results across repeated screening rounds. We summarize MCED false-positive burden as the mean number of false positives per 100 individuals.

### 1.6 Natural history assumptions

Natural history assumptions for CRC were based on the calibrated CRC-SPIN model. For the additional MCED-targeted cancers, we considered five alternative sets of natural history inputs (Supplemental Table 1). Input sets A-C reflect plausible detectable preclinical windows for currently available MCED assays, informed by analyses of stored blood samples suggesting that MCED test positivity is minimal beyond approximately two years prior to clinical diagnosis.^30, 31^ Input sets D and E are based on literature-derived estimates of cancer-specific preclinical durations obtained from expert opinion or natural history models calibrated to screening trial data and reflect optimistic assumptions about the MCED detectability window, assuming that the cell-free DNA signal has a detectable sojourn time similar to other (mostly imaging-based) modalities.

For the literature-based input sets, OMST values for lung, pancreas, and ovarian cancers were derived as the median of estimates from published modeling studies Supplemental Table 2. Stomach and esophageal cancers were assigned an OMST of 5 years based on estimates from Luebeck et al.,^32^ and cancers without specific published estimates (anus, bladder, liver, lymphoma, and head and neck) were assumed to have an OMST of 3 years. LMST was set to either 0.5 or 1.0 years across all cancer sites. Full details for the literature-based input sets are provided in Supplemental Table 3.

### 1.7 Sensitivity analysis

The stage-shift model assumes that screen-detected cancers have the same post–lead-time survival as clinically detected cancers of the same stage. Because MCEDsim collapses AJCC-7 stages into broad early and late categories, this assumption may underestimate the mortality benefit of screening. To represent a more optimistic assumption about screening benefit, we assigned post–lead-time survival for screen-detected, MCED-targeted non-CRC cancers using SEER 17 (2010–2021) Stage 0–I survival for early-stage disease and Stage III survival for late-stage disease. For pancreatic cancer, early-stage survival was likewise based on Stage 0–I, but late-stage survival was based on Stage II.

## 2 Results

### 2.1 Calibration of MCEDsim

Observed and predicted clinical incidence rates (Supplemental Figures 4 and 5) indicate that the MCEDsim natural history models closely reproduce SEER-observed incidence rates across sex, site, age, and stage for the majority of targeted cancer sites. The fitted models demonstrate some deviations from observed incidence for lymphoma, as well as cancer sites where the incidence decreases at the oldest ages, such as ovarian and uterine. The fitted incidence data were virtually identical across natural history inputs.

Supplemental Figure 6 shows observed and predicted survival curves for non-CRC MCED-targeted cancers by sex, cancer site, and stage under baseline and optimistic assumptions. Supplemental Figure 7 shows the observed and predicted survival curves by stage 0-IV for CRC. The log-logistic models demonstrated good fit to the observed Kaplan-Meier survival curves across cancer sites, stages, and sex, with predicted survival probabilities closely tracking observed patterns over the 15-year follow-up period.

### 2.2 CRC and MCED-targeted cancer incidence

Lifetime cumulative CRC incidence was substantially reduced under colonoscopy and FIT-annual strategies relative to the Control strategy (lifetime cumulative incidence control: 10%; colonoscopy: 0.5%; FIT-annual: 1%). In contrast, MCED strategies screening strategies showed no reduction in CRC incidence. Total all-cancer lifetime cumulative incidence followed the same pattern (Control: 34–35%; Colonoscopy: 25%; FIT-annual: 25–26%; MCED strategies: 34–35%; ranges reflect natural history assumptions A–E).

### 2.3 Cumulative late-stage disease

Table 4 and Figure 1 summarize lifetime cumulative incidence of cancer diagnosed in late-stage for all cancers and CRC specifically across screening strategies and natural history assumptions. In the Control strategy, 18% of individuals experienced a late-stage cancer diagnosis during their lifetime. Compared to the Control strategy, Colonoscopy and FIT-annual were associated with 26% and 25% relative reductions in overall late-stage diagnoses, respectively. MCED annual reduced late-stage disease by 20–32%, with the more optimistic literature-informed natural history assumptions D and E achieving larger reductions. Less frequent MCED screening yielded substantially smaller benefits: MCED biennial screening reduced late-stage disease by 9–12%, and MCED triennial by 6–7%.

**Figure 1:**
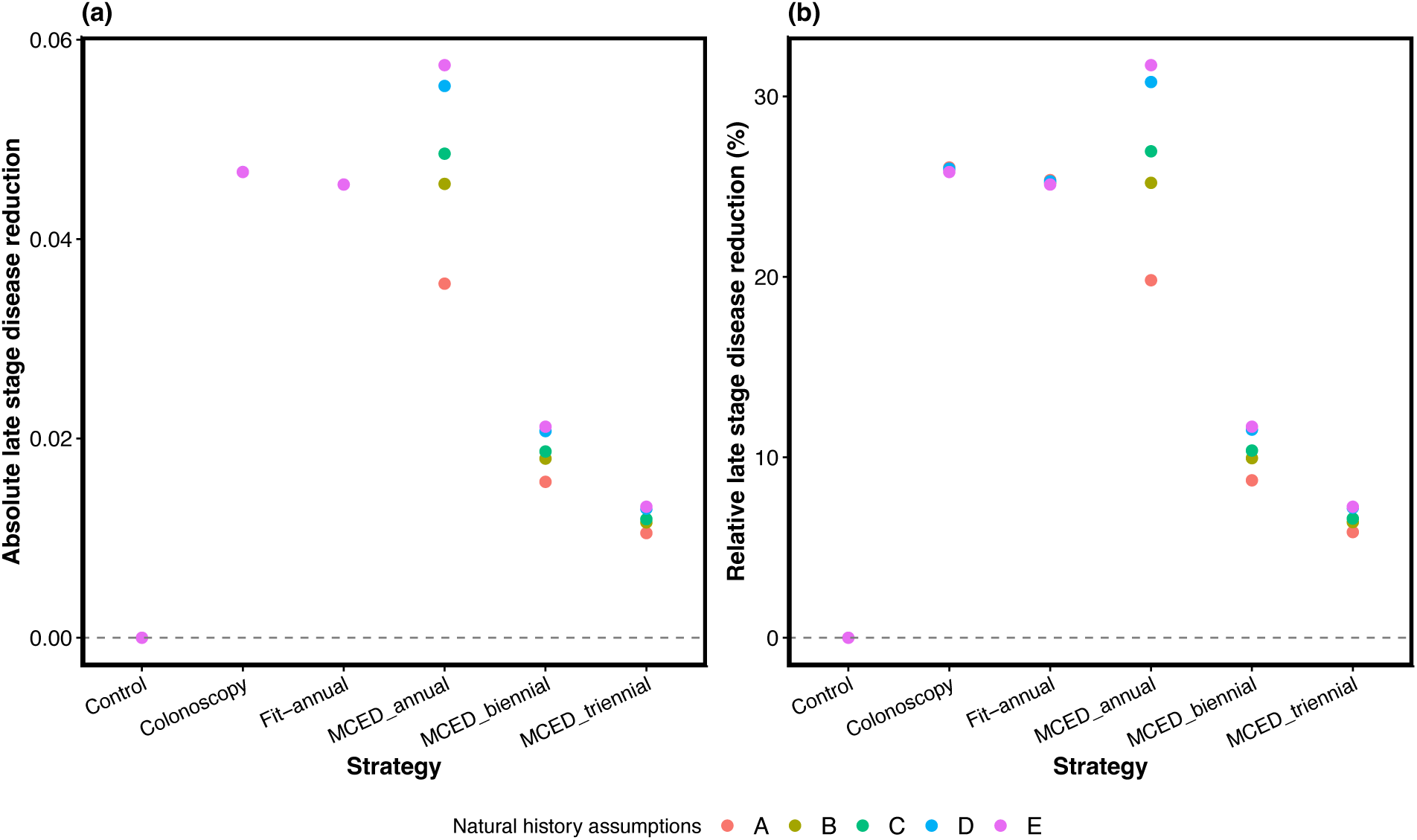
Absolute and relative late-stage disease reduction across screening strategies and natural history assumptions.

**Table 4:** Late-stage cancer outcomes under different natural assumptions and screening strategies. Abbreviations: NH=natural history; LS=late stage.

| NH assumption | Strategy | Percent LS (all) | Relative reduction (%) (all) | Percent LS (CRC) | Relative reduction (%) (CRC) |
| --- | --- | --- | --- | --- | --- |
| A | Control | 17.9 | 0.0 | 4.9 | 0.0 |
| B | Control | 18.1 | 0.0 | 4.9 | 0.0 |
| C | Control | 18.1 | 0.0 | 4.9 | 0.0 |
| D | Control | 18.0 | 0.0 | 4.9 | 0.0 |
| E | Control | 18.1 | 0.0 | 4.9 | 0.0 |
| A | Colonoscopy | 13.3 | 26.1 | 0.2 | 96.4 |
| B | Colonoscopy | 13.4 | 25.9 | 0.2 | 96.4 |
| C | Colonoscopy | 13.4 | 25.8 | 0.2 | 96.4 |
| D | Colonoscopy | 13.3 | 26.0 | 0.2 | 96.4 |
| E | Colonoscopy | 13.5 | 25.8 | 0.2 | 96.4 |
| A | Fit-annual | 13.4 | 25.4 | 0.3 | 93.9 |
| B | Fit-annual | 13.5 | 25.2 | 0.3 | 93.9 |
| C | Fit-annual | 13.5 | 25.2 | 0.3 | 93.9 |
| D | Fit-annual | 13.5 | 25.3 | 0.3 | 93.9 |
| E | Fit-annual | 13.6 | 25.1 | 0.3 | 93.9 |
| A | MCED_annual | 14.4 | 19.7 | 2.9 | 40.3 |
| B | MCED_annual | 13.5 | 25.2 | 2.9 | 40.3 |
| C | MCED_annual | 13.2 | 27.1 | 2.9 | 40.3 |
| D | MCED_annual | 12.4 | 31.0 | 2.9 | 40.3 |
| E | MCED_annual | 12.4 | 31.8 | 2.9 | 40.3 |
| A | MCED_biennial | 16.4 | 8.7 | 3.6 | 26.7 |
| B | MCED_biennial | 16.3 | 9.9 | 3.6 | 26.7 |
| C | MCED_biennial | 16.2 | 10.4 | 3.6 | 26.7 |
| D | MCED_biennial | 15.9 | 11.6 | 3.6 | 26.7 |
| E | MCED_biennial | 16.0 | 11.7 | 3.6 | 26.7 |
| A | MCED_triennial | 16.9 | 5.8 | 3.9 | 19.2 |
| B | MCED_triennial | 16.9 | 6.3 | 3.9 | 19.1 |
| C | MCED_triennial | 16.9 | 6.6 | 3.9 | 19.1 |
| D | MCED_triennial | 16.7 | 7.1 | 3.9 | 19.1 |
| E | MCED_triennial | 16.8 | 7.2 | 3.9 | 19.1 |

For CRC specifically, we observed a 5% lifetime incidence of late-stage diagnoses in the Control strategy. Colonoscopy and FIT-annual strategies were associated with a relative reduction of 96% and 94%, respectively. MCED strategies were associated with 19%–40% reductions, with larger decreases for more frequent testing schedules.

### 2.4 Life years gained

Across all assumptions, guideline-recommended CRC screening strategies yielded larger life-year gains than any MCED screening strategy (Figure 2, Table 5). Colonoscopy produced the greatest gains, increasing life expectancy by 0.70 years (relative increase 0.9%), followed by FIT annual with gains of 0.65 life-years (relative increase 0.8%). MCED annual screening yielded 33%–46% of the LYG of colonoscopy; MCED biennial screening, 19%–23%; and MCED triennial, 13%-16%. LYG from MCED screening increased progressively from assumptions A to E, consistent with longer early-stage detectable preclinical periods yielding greater opportunities for stage shift and mortality reduction.

**Figure 2:**
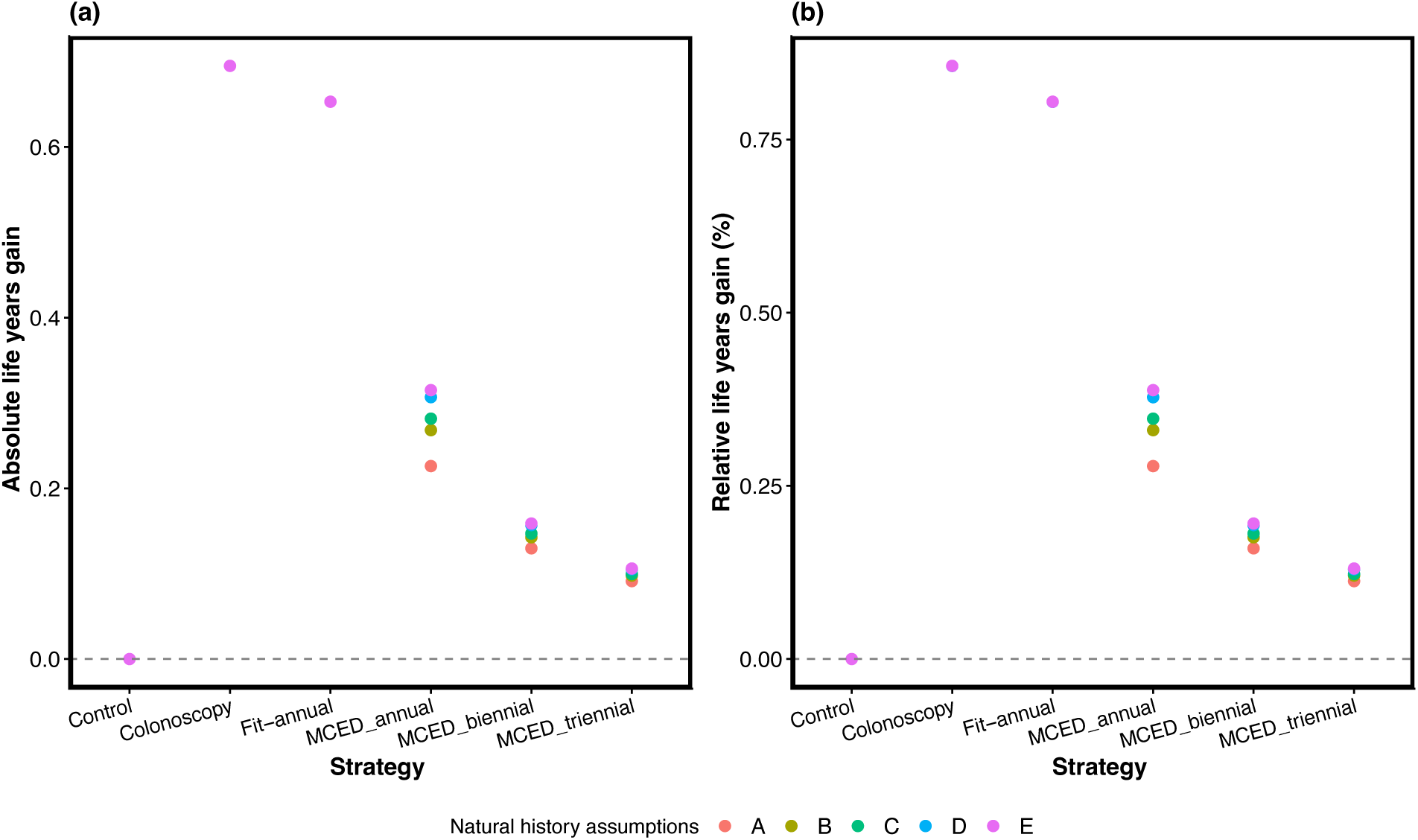
Absolute and relative life years gain across screening strategies under base case survival assumptions.

**Table 5:**
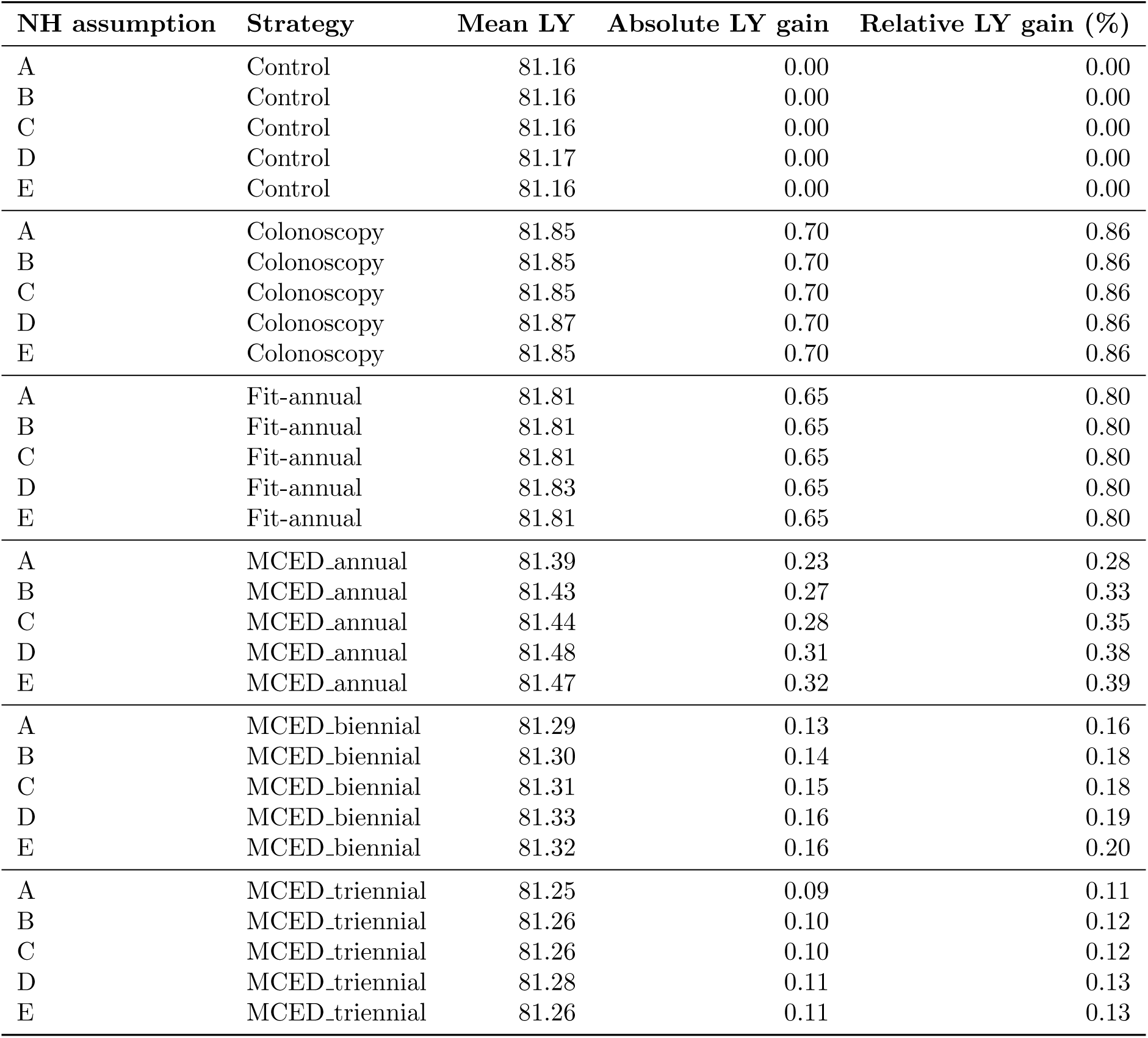
Life-year outcomes under different natural history assumptions and screening strategies. Cancer-specific death is generated according to the base-case assumptions. Abbreviations: NH=natural history; LY=life-years.

### 2.5 Sensitivity analysis

Under the more optimistic survival assumptions described in Section 1.7, the estimated benefits of increased across all MCED strategies (Supplemental Table 4). The largest effect was observed for annual screening, where absolute life-year gains increased from 0.23–0.32 years to 0.26–0.36 years from baseline to optimistic assumptions. Smaller increases were observed for biennial and triennial screening. Notably, despite improved life years under the optimistic survival assumptions, guideline-recommended CRC screening still outperformed the MCED-only strategies: MCED annual was associated with 37%-51% of the LYG of colonoscopy.

### 2.6 MCED false positive burden

False positive rates for MCED strategies were driven by screening frequency, with natural history assumptions contributing only minor variation across strategies. MCED annual screening generated approximately 14 false positives per 100 individuals, compared with 7 for MCED biennial and 5 for MCED triennial.

### 2.7 Contribution of CRC to disease burden and MCED screening benefit

In the Control strategy, CRC accounted for 29% of the cumulative lifetime incidence of the 13 cancer sites considered in our model, 27% of late-stage diagnoses, and 33% of cancer-specific deaths.

Granular breakdowns of MCED screening benefit by CRC and non-CRC cancers are provided in Supplemental Tables 5, 6, and 7. CRC made a disproportionately large contribution to the overall benefit of MCED screening (Table 6). Detection of CRC accounted for 34%–55% of the absolute reduction in late-stage disease across models A–E. Under less frequent screening schedules, this proportion increased. The pattern was even more pronounced for LYG, with CRC accounting for 50%–70% under annual screening and up to 93% under triennial screening.

**Table 6:** Fraction of total MCED screening benefit attributable to colorectal cancer.

| Outcome | Screening interval | Natural history assumption |  |  |  |  |
| --- | --- | --- | --- | --- | --- | --- |
|  |  | A | B | C | D | E |
| Absolute late-stage difference | Annual | 55.3% | 43.0% | 40.0% | 35.0% | 34.0% |
|  | Biennial | 83.5% | 72.6% | 69.0% | 62.2% | 61.2% |
|  | Triennial | 88.9% | 81.4% | 78.4% | 72.5% | 71.0% |
| Life-years gained | Annual | 70.2% | 59.4% | 56.1% | 51.6% | 50.1% |
|  | Biennial | 88.8% | 81.2% | 77.8% | 73.3% | 71.9% |
|  | Triennial | 92.8% | 87.0% | 84.6% | 81.0% | 79.7% |

## 3 Discussion

In this microsimulation study, we found that guideline-recommended CRC screening strategies, including colonoscopy and annual FIT, generated more life-years gained than MCED-only screening strategies under optimistic assumptions regarding preclinical sensitivity of a currently available MCED test.^15^ This pattern was consistent across a broad range of assumptions regarding preclinical disease duration and the survival benefits associated with earlier cancer detection. These findings indicate that replacing guideline-recommended CRC screening with current MCED testing would likely reduce population-level life expectancy despite the ability of MCED assays to detect multiple cancer types.

These results are not entirely surprising in light of CRC’s characteristics. Of the cancer sites modeled, CRC accounted for 29% of lifetime cancer incidence and 33% of cancer deaths. Thus, the ability of FIT and colonoscopy to reduce CRC incidence through adenoma detection and removal yields substantial gains in life expectancy. By contrast, MCED assays detect CRC but, given their high specificity and methylation-based signal, are expected to have negligible sensitivity for precursor lesions, and thus provide no equivalent prevention of incident CRC.

Even beyond prevention, CRC is a particularly favorable target for early detection. The long natural history of CRC^33^ provides ample opportunity for screen detection, while favorable survival differences between early- and late-stage disease allow stage shifts to translate into meaningful life-years gained. These characteristics help explain the large contribution of CRC to MCED screening benefit in addition to explaining a portion of the benefit of guideline-recommended strategies.

Overall, our findings underscore the importance of public health policies and messaging that continue to encourage guideline-recommended CRC screening, and to discourage perceptions that MCED screening may act as a substitute. Nonetheless, MCED strategies may provide meaningful incremental benefit when added to, rather than substituted for, guideline-recommended CRC screening. Indeed, our decomposition analyses suggest that the non-CRC benefits of MCED screening might contribute an additional 13–36% of the life-years gained from colonoscopy screening alone. Future incarnations of MCED tests with higher early-stage sensitivity have the potential to yield greater marginal benefit.

Our approach has several limitations, including uncertainty in the natural history of cancers targeted by MCED tests and their detectability through cell-free DNA methylation signals. Although we explored a range of natural history models, we did not consider the possibility that CRC has a shorter preclinical detectable window for cell-free DNA methylation signals than for FIT or colonoscopy. To the extent that this is true, the contribution of CRC to MCED screening benefit may be overstated. Survival assumptions also represent a limitation. Although our parametric models generally fit the observed data well, extrapolations beyond the available data are uncertain. Survival curves were based on cases diagnosed between 2010 and 2015 and may not reflect current or future treatment advances. In addition, CRC survival from this time frame may include lead-time bias from screen-detected cancers, potentially overstating the contribution of CRC to MCED screening benefit. Importantly, our analysis focused on an idealized implementation of each strategy, assuming perfect adherence to screening and complete diagnostic follow-up. Finally, we focused primarily on screening benefits and did not undertake a formal cost-effectiveness analysis or comprehensive evaluation of harms associated with either MCED or guideline-recommended CRC screening.

## 4 Conclusion

In conclusion, this simulation study suggests that currently available MCED tests are unlikely to be effective replacements for guideline-recommended CRC screening, largely because they lack the preventive benefit associated with precuros lesion detection and removal. MCED tests may provide additional benefit when used alongside existing screening programs, but should not be viewed as substitutes for established CRC screening.

## 5 Author contributions

Conceptualization: IA, JL, CR. Methodology: IA, JL, CR, CM. Supervision: JL, CR. Analysis: IA, JL. Writing–original draft: IA, JL. Writing–review and editing: IA, JL, CR, CM.

## 6 Funding acknowledgements

IA and JL were funded by Cancer Early Detection Advanced Research Center grants Full 2024-1960. CR and CM were supported by U01 CA253913 from the National Cancer Institute as part of the Cancer Intervention and Surveillance Modeling Network (CISNET). Its contents are solely the responsibility of the authors and do not necessarily represent the official views of the National Cancer Institute.

## Data Availability

The model code and data for MCEDsim are available on Github.

https://github.com/Ishfaq8899/March_MCEDsim_Carolyn

