## Supplementary Tables and Figures for "Should Multi-Cancer Early Detection Testing Replace Guideline-Recommended Colorectal Cancer Screening? A Comparative Modeling Analysis"

### A. Supplementary Tables

Supplemental Table 1: Alternative natural history assumptions for non-CRC MCED-targeted cancers. Models A–C reflect plausible natural history assumptions based on stored-blood studies. Literature-based natural history models (D–E) reflect published estimates of cancer-specific preclinical durations derived from expert opinion and prior screening trial data

| **Natural history assumptions** | **OMST (years)** | **LMST (years)** | EMST (years) |
|---|---|---|---|
| A | 1.0 | 0.5 | 0.75 |
| B | 2.0 | 1.0 | 1.51 |
| C | 2.0 | 0.5 | 1.75 |
| D (Supplemental Table 3) | | 1.0 | |
| E (Supplemental Table 3) | | 0.5 | |

Supplemental Table 2: Literature-based estimates of overall mean sojourn time (OMST) by cancer site.

| **Cancer Site** | **MST (years)** | **Trial or Study** | **References** |
|---|---|---|---|
| **Lung** | 3.09–5.32 (men); 3.35–6.01 (women) | NLST; PLCO | Ten Haaf et al.[?] |
| | 1.8 | Mayo Lung Screening Trial | Pinsky[?] |
| | 3.6 (3–4.3) | NLST | Patz et al.[?] |
| | 2.06 (0.42–3.83) | 6-study summary | Chien & Chen[?] |
| | 2.24 (1.57–3.35) | Mayo Lung Project | Wu et al.[?] |
| **Colorectal** | 4 (2–5) | SimCRC | – |
| | 3.6 (2–5) | CRC-SPIN | – |
| | 4.7 (1–7) | MISCAN | – |
| | 4.5–5.8 | German Screening Registry | Brenner et al.[?] |
| **Pancreas** | 3 | – | Luebeck et al.[?] |
| | 2.5–3 | – | Sharma et al.[?] |
| **Ovary** | 2 (1.8–2.1) | – | Ishizawa et al.[?] |

Supplemental Table 3: Literature-based natural history assumptions (D-E) for MCED-targeted cancers.

| **Cancer Site** | **OMST (years)** | **LMST (years)** | **Source** |
|---|---|---|---|
| Anus | 3.0 | 0.5 / 1.0 | Supplemental Table 2 |
| Bladder | 3.0 | 0.5 / 1.0 | Assumed |
| Esophagus | 5.0 | 0.5 / 1.0 | Luebeck et al.[?] |
| Gastric | 5.0 | 0.5 / 1.0 | Luebeck et al.[?] |
| Headandneck | 3.0 | 0.5 / 1.0 | Assumed |
| Liver | 3.0 | 0.5 / 1.0 | Assumed |
| Lung | 3.1 | 0.5 / 1.0 | Supplemental Table 2 |
| Lymphoma | 3.0 | 0.5 / 1.0 | Assumed |
| Ovary | 2.0 | 0.5 / 1.0 | Supplemental Table 2 |
| Pancreas | 2.9 | 0.5 / 1.0 | Supplemental Table 2 |
| Renal | 3 | 0.5 / 1.0 | Assumed |
| Uterine | 3 | 0.5 / 1.0 | Assumed |

Supplemental Table 4: Sensitivity analysis: life-year outcomes under different natural history assumptions and screening strategies. Cancer-specific death is generated according to the optimistic assumptions. Abbreviations: NH=natural history; LY=life-years.

| NH assumption | Strategy | Mean LY | Absolute LY gain | Relative LY gain (%) |
|---|---|---|---|---|
| A | MCED_annual | 81.42 | 0.26 | 0.32 |
| B | MCED_annual | 81.48 | 0.32 | 0.39 |
| C | MCED_annual | 81.48 | 0.32 | 0.40 |
| D | MCED_annual | 81.53 | 0.36 | 0.44 |
| E | MCED_annual | 81.52 | 0.36 | 0.44 |
| A | MCED_biennial | 81.30 | 0.14 | 0.17 |
| B | MCED_biennial | 81.32 | 0.16 | 0.19 |
| C | MCED_biennial | 81.32 | 0.16 | 0.19 |
| D | MCED_biennial | 81.34 | 0.17 | 0.21 |
| E | MCED_biennial | 81.33 | 0.17 | 0.21 |
| A | MCED_triennial | 81.25 | 0.09 | 0.12 |
| B | MCED_triennial | 81.26 | 0.10 | 0.13 |
| C | MCED_triennial | 81.26 | 0.10 | 0.13 |
| D | MCED_triennial | 81.28 | 0.11 | 0.14 |
| E | MCED_triennial | 81.27 | 0.11 | 0.14 |

Supplemental Table 5: Decomposition of non-CRC and CRC contributions to MCED life-year outcomes under different natural history assumptions. Cancer-specific death is generated according to the base case assumptions. Abbreviations: NH=natural history; LY=life-years.

| NH assumption | Strategy | Mean LY | Absolute LY gain | Relative LY gain (%) |
|---|---|---|---|---|
| A | MCED_annual (non-CRC) | 81.23 | 0.07 | 0.09 |
| B | MCED_annual (non-CRC) | 81.27 | 0.11 | 0.14 |
| C | MCED_annual (non-CRC) | 81.28 | 0.13 | 0.15 |
| D | MCED_annual (non-CRC) | 81.32 | 0.15 | 0.19 |
| E | MCED_annual (non-CRC) | 81.32 | 0.16 | 0.20 |
| A | MCED_biennial (non-CRC) | 81.17 | 0.02 | 0.02 |
| B | MCED_biennial (non-CRC) | 81.19 | 0.03 | 0.03 |
| C | MCED_biennial (non-CRC) | 81.19 | 0.03 | 0.04 |
| D | MCED_biennial (non-CRC) | 81.22 | 0.04 | 0.05 |
| E | MCED_biennial (non- CRC) | 81.20 | 0.04 | 0.05 |
| A | MCED_triennial (non-CRC) | 81.17 | 0.01 | 0.01 |
| B | MCED_triennial (non-CRC) | 81.17 | 0.01 | 0.02 |
| C | MCED_triennial (non-CRC) | 81.17 | 0.01 | 0.02 |
| D | MCED_triennial (non-CRC) | 81.19 | 0.02 | 0.03 |
| E | MCED_triennial (non-CRC) | 81.18 | 0.02 | 0.03 |
| A | MCED_annual (CRC only) | 81.32 | 0.16 | 0.19 |
| B | MCED_annual (CRC only) | 81.32 | 0.16 | 0.19 |
| C | MCED_annual (CRC only) | 81.31 | 0.16 | 0.19 |
| D | MCED_annual (CRC only) | 81.33 | 0.16 | 0.19 |
| E | MCED_annual (CRC only) | 81.31 | 0.16 | 0.19 |
| A | MCED_biennial (CRC only) | 81.27 | 0.11 | 0.14 |
| B | MCED_biennial (CRC only) | 81.27 | 0.11 | 0.14 |
| C | MCED_biennial (CRC only) | 81.27 | 0.11 | 0.14 |
| D | MCED_biennial (CRC only) | 81.29 | 0.11 | 0.14 |
| E | MCED_biennial (CRC only) | 81.27 | 0.11 | 0.14 |
| A | MCED_triennial (CRC only) | 81.24 | 0.08 | 0.10 |
| B | MCED_triennial (CRC only) | 81.24 | 0.08 | 0.10 |
| C | MCED_triennial (CRC only) | 81.24 | 0.08 | 0.10 |
| D | MCED_triennial (CRC only) | 81.26 | 0.08 | 0.10 |
| E | MCED_triennial (CRC only) | 81.24 | 0.08 | 0.10 |

Supplemental Table 6: Decomposition of non-CRC and CRC contributions to MCED life-year outcomes under different natural history assumptions. Cancer-specific death is generated according to the optimistic assumptions. Abbreviations: NH=natural history; LY=life-years.

| NH assumption | Strategy | Mean LY | Absolute LY gain | Relative LY gain (%) |
|---|---|---|---|---|
| A | MCED_annual (no-CRC) | 81.27 | 0.11 | 0.13 |
| B | MCED_annual (no-CRC) | 81.32 | 0.16 | 0.20 |
| C | MCED_annual (no-CRC) | 81.33 | 0.17 | 0.21 |
| D | MCED_annual (no-CRC) | 81.37 | 0.20 | 0.25 |
| E | MCED_annual (no-CRC) | 81.36 | 0.20 | 0.25 |
| A | MCED_biennial (no-CRC) | 81.18 | 0.02 | 0.03 |
| B | MCED_biennial (no-CRC) | 81.20 | 0.04 | 0.05 |
| C | MCED_biennial (no-CRC) | 81.20 | 0.04 | 0.05 |
| D | MCED_biennial (no-CRC) | 81.23 | 0.06 | 0.07 |
| E | MCED_biennial (no-CRC) | 81.21 | 0.06 | 0.07 |
| A | MCED_triennial (no-CRC) | 81.17 | 0.01 | 0.01 |
| B | MCED_triennial (no-CRC) | 81.18 | 0.02 | 0.02 |
| C | MCED_triennial (no-CRC) | 81.18 | 0.02 | 0.02 |
| D | MCED_triennial (no-CRC) | 81.20 | 0.03 | 0.03 |
| E | MCED_triennial (no-CRC) | 81.18 | 0.03 | 0.03 |

Supplemental Table 7: Decomposition of non-CRC and CRC contributions to MCED late-stage disease outcomes under different natural history assumptions. Abbreviations: NH=natural history; LY=life-years.

| NH assumption | Strategy | Percent LS (all) | Relative reduction (%) (all) | Percent LS (CRC) | Relative reduction (%) (CRC) |
|---|---|---|---|---|---|
| A | MCED_annual (non-CRC) | 16.4 | 8.8 | 4.9 | 0.0 |
| B | MCED_annual (non-CRC) | 15.5 | 14.4 | 4.9 | 0.0 |
| C | MCED_annual (non-CRC) | 15.2 | 16.3 | 4.9 | 0.0 |
| D | MCED_annual (non-CRC) | 14.4 | 20.2 | 4.9 | 0.0 |
| E | MCED_annual (non-CRC) | 14.3 | 21.0 | 4.9 | 0.0 |
| A | MCED_biennial (non-CRC) | 17.7 | 1.5 | 4.9 | 0.0 |
| B | MCED_biennial (non-CRC) | 17.6 | 2.7 | 4.9 | 0.0 |
| C | MCED_biennial (non-CRC) | 17.5 | 3.2 | 4.9 | 0.0 |
| D | MCED_biennial (non-CRC) | 17.2 | 4.4 | 4.9 | 0.0 |
| E | MCED_biennial (non-CRC) | 17.3 | 4.6 | 4.9 | 0.0 |
| A | MCED_triennial (non-CRC) | 17.8 | 0.7 | 4.9 | 0.0 |
| B | MCED_triennial (non-CRC) | 17.9 | 1.2 | 4.9 | 0.0 |
| C | MCED_triennial (non-CRC) | 17.8 | 1.4 | 4.9 | 0.0 |
| D | MCED_triennial (non-CRC) | 17.7 | 2.0 | 4.9 | 0.0 |
| E | MCED_triennial (non-CRC) | 17.8 | 2.1 | 4.9 | 0.0 |
| A | MCED_annual (CRC only) | 16.0 | 10.9 | 2.9 | 40.3 |
| B | MCED_annual (CRC only) | 16.1 | 10.8 | 2.9 | 40.3 |
| C | MCED_annual (CRC only) | 16.1 | 10.8 | 2.9 | 40.3 |
| D | MCED_annual (CRC only) | 16.1 | 10.9 | 2.9 | 40.3 |
| E | MCED_annual (CRC only) | 16.2 | 10.8 | 2.9 | 40.3 |
| A | MCED_biennial (CRC only) | 16.6 | 7.3 | 3.6 | 26.7 |
| B | MCED_biennial (CRC only) | 16.8 | 7.2 | 3.6 | 26.7 |
| C | MCED_biennial (CRC only) | 16.8 | 7.2 | 3.6 | 26.7 |
| D | MCED_biennial (CRC only) | 16.7 | 7.2 | 3.6 | 26.7 |
| E | MCED_biennial (CRC only) | 16.8 | 7.2 | 3.6 | 26.7 |
| A | MCED_triennial (CRC only) | 17.0 | 5.2 | 3.9 | 19.2 |
| B | MCED_triennial (CRC only) | 17.1 | 5.1 | 3.9 | 19.1 |
| C | MCED_triennial (CRC only) | 17.2 | 5.1 | 3.9 | 19.1 |
| D | MCED_triennial (CRC only) | 17.1 | 5.2 | 3.9 | 19.1 |
| E | MCED_triennial (CRC only) | 17.2 | 5.1 | 3.9 | 19.1 |

### B. Supplementary Figures

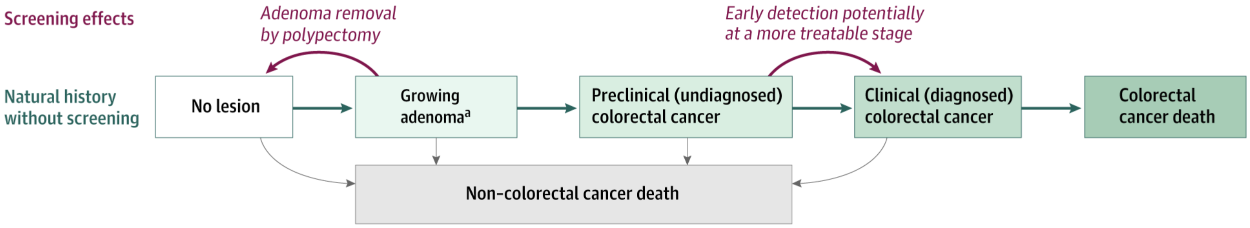

Supplemental Figure 1: Schematic of CRC-SPIN natural history model.

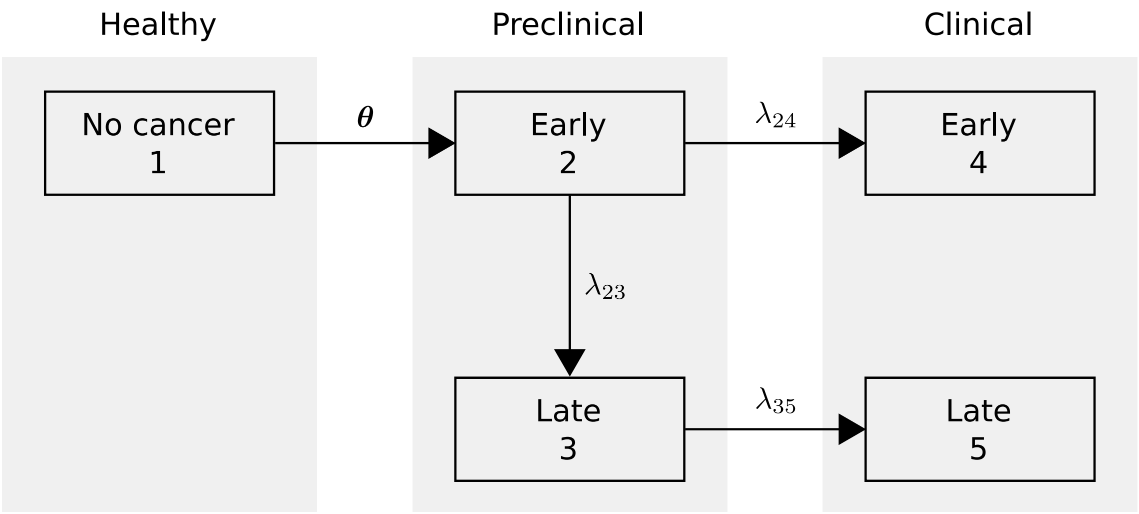

Supplemental Figure 2: Schematic of MCEDsim five-state natural history model applied across non-CRC cancer sites.

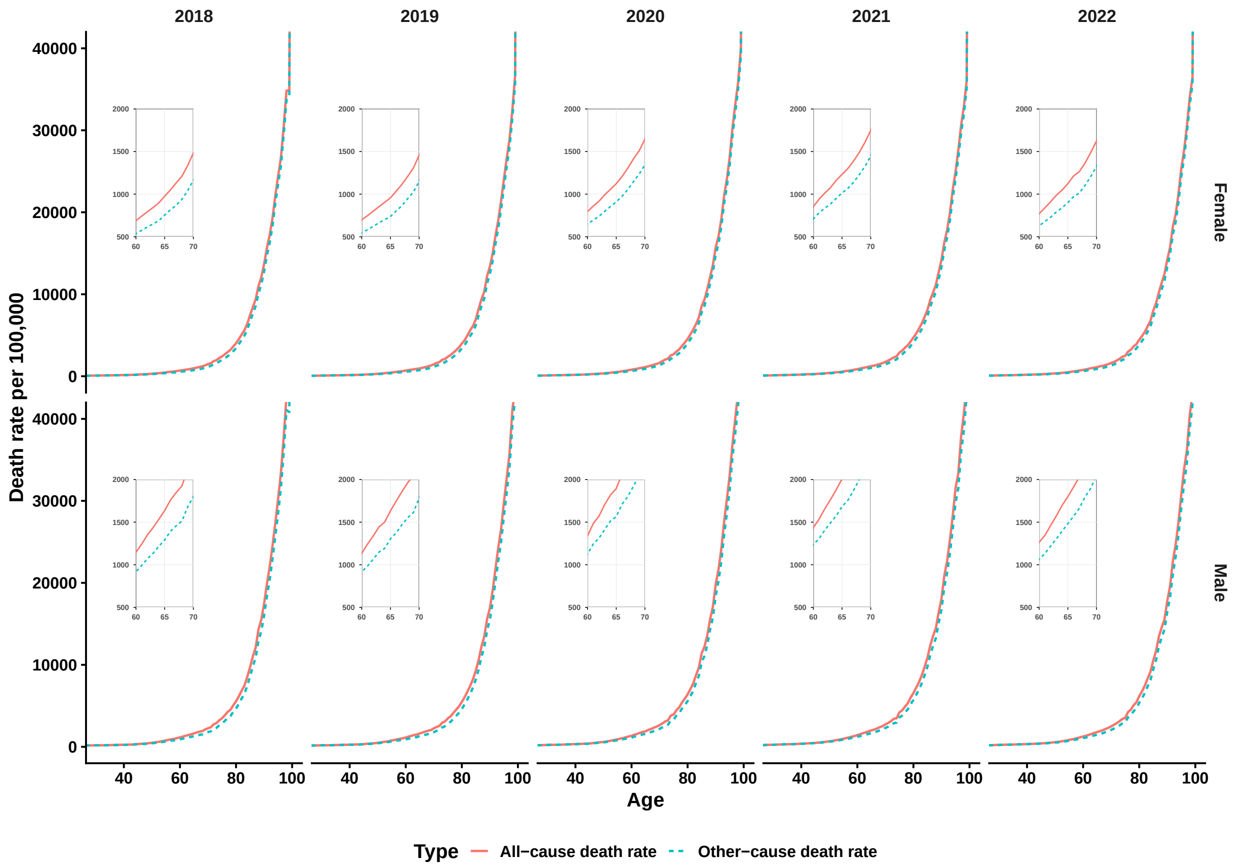

Supplemental Figure 3: Comparison of all cause mortality (solid red) and other cause mortality excluding MCED targeted cancers (dashed blue) in the US, 2018-2022. Main panels show death rates across ages 40-100 years stratified by sex. Inset panels display zoomed views for ages 60-70 years across individual years (2018-2022) to more clearly visualize differences between all-cause and other-cause mortality rates.

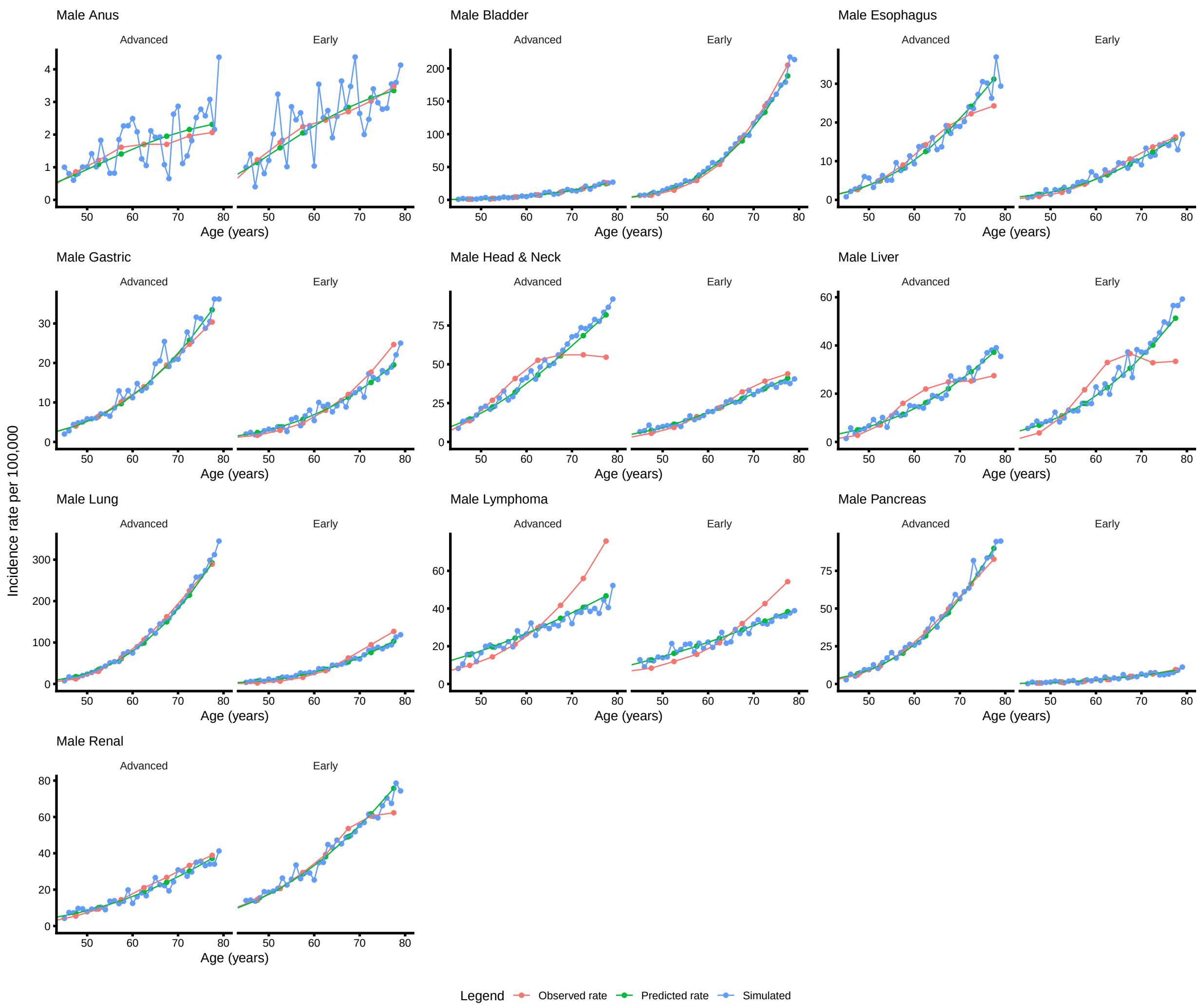

Supplemental Figure 4: Comparison of observed cancer incidence rates (red) against predicted (green), and simulated (blue) for males. Age-specific incidence rates per 100,000 person-years are shown for each cancer site, stratified by early and late stage.

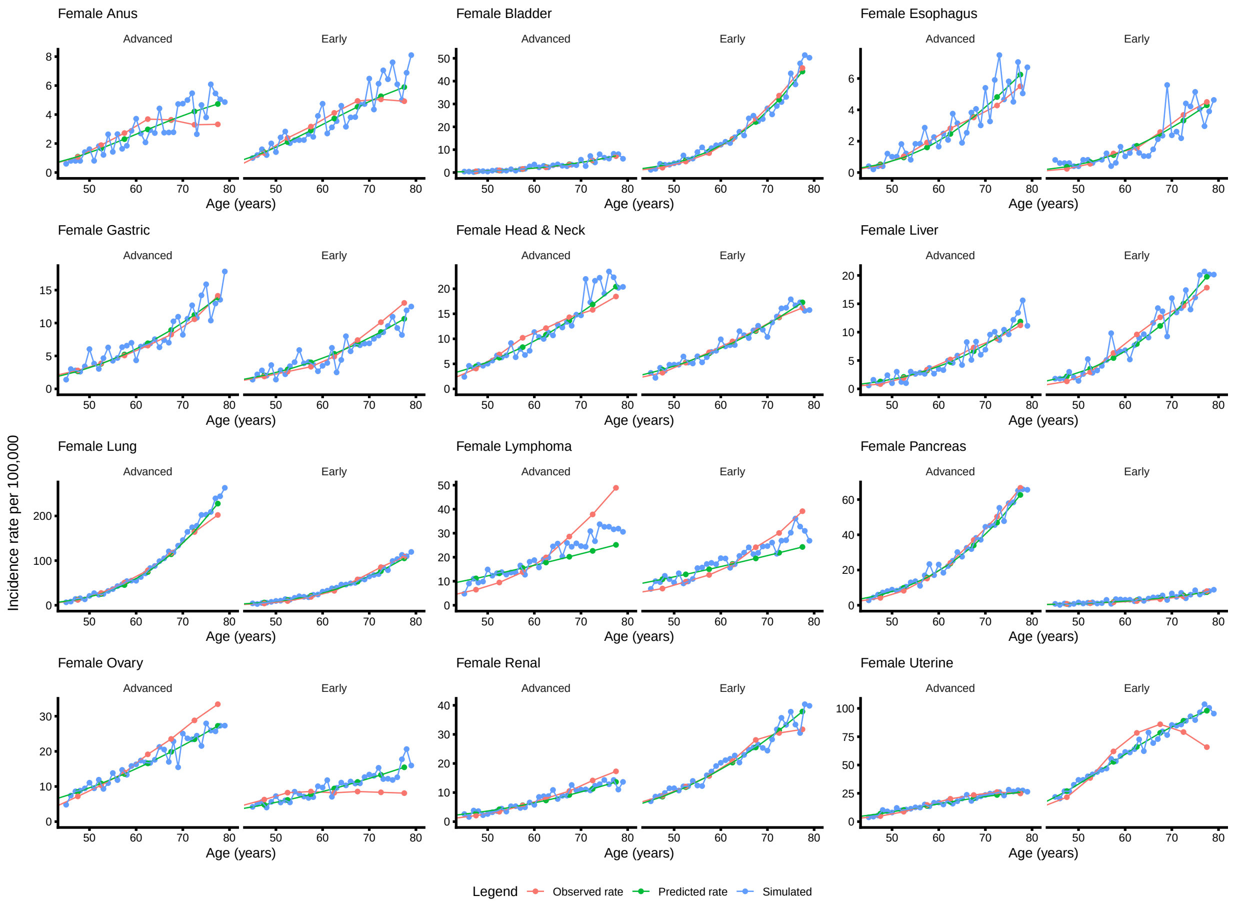

Supplemental Figure 5: Comparison of observed cancer incidence rates (red) against predicted (green), and simulated (blue) for females. Age-specific incidence rates per 100,000 person-years are shown for each cancer site, stratified by early and late stage.

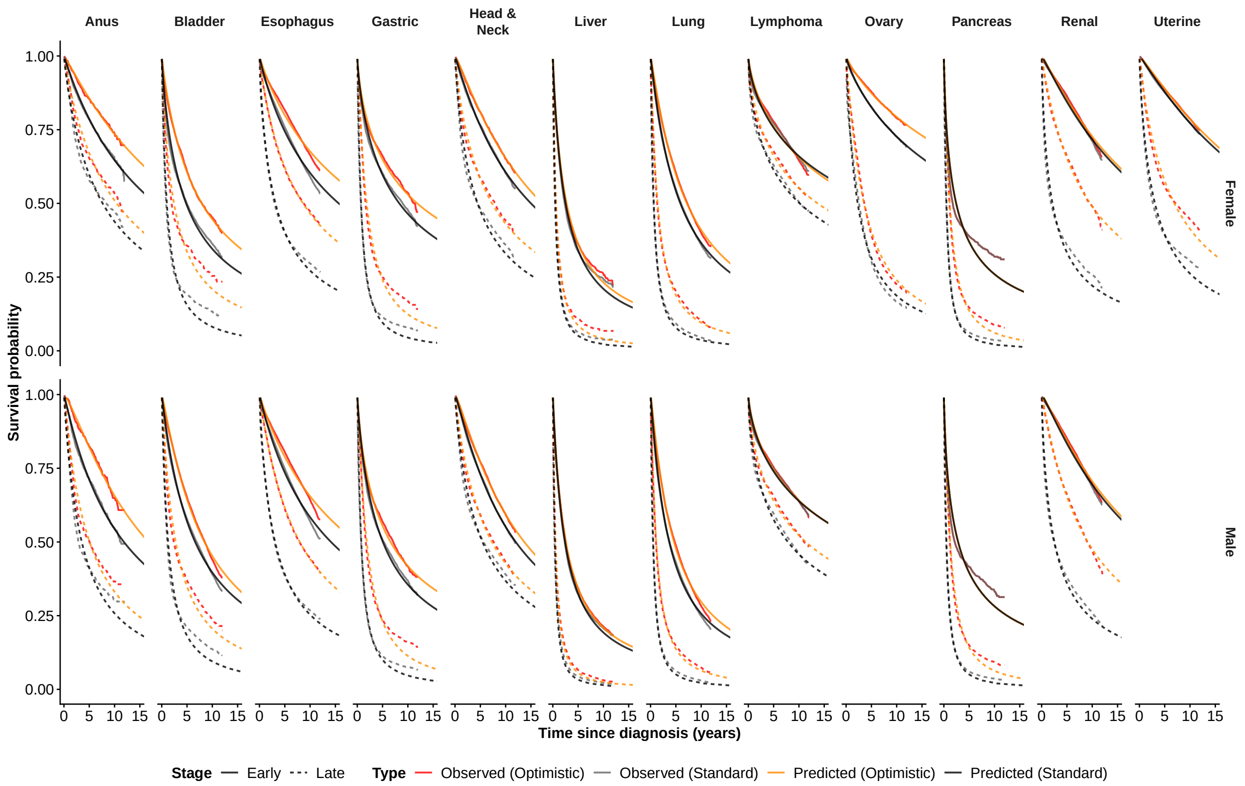

Supplemental Figure 6: Observed and predicted survival curves for non-CRC MCED-targeted cancers under base case and optimistic stage groupings, by sex, cancer site, and stage. Kaplan-Meier survival curves ("Observed") estimated from SEER cases diagnosed between 2010–2015 are compared with predicted curves ("Predicted") from fitted log-logistic parametric survival models.

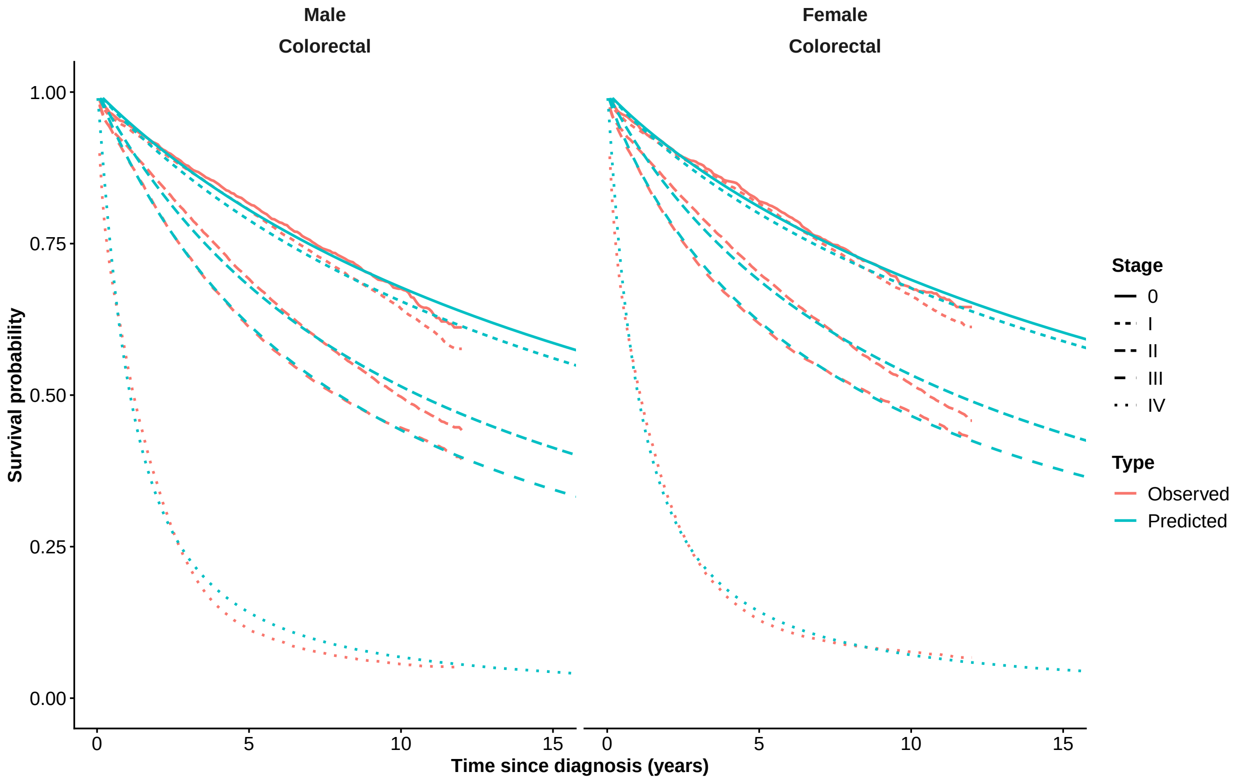

Supplemental Figure 7: Observed and predicted survival curves for CRC by sex, cancer site, and AJCC-7 stage O-IV. Kaplan-Meier survival curves ("Observed") estimated from SEER cases diagnosed between 2010–2015 are compared with predicted curves ("Predicted") from fitted log-logistic parametric survival models.

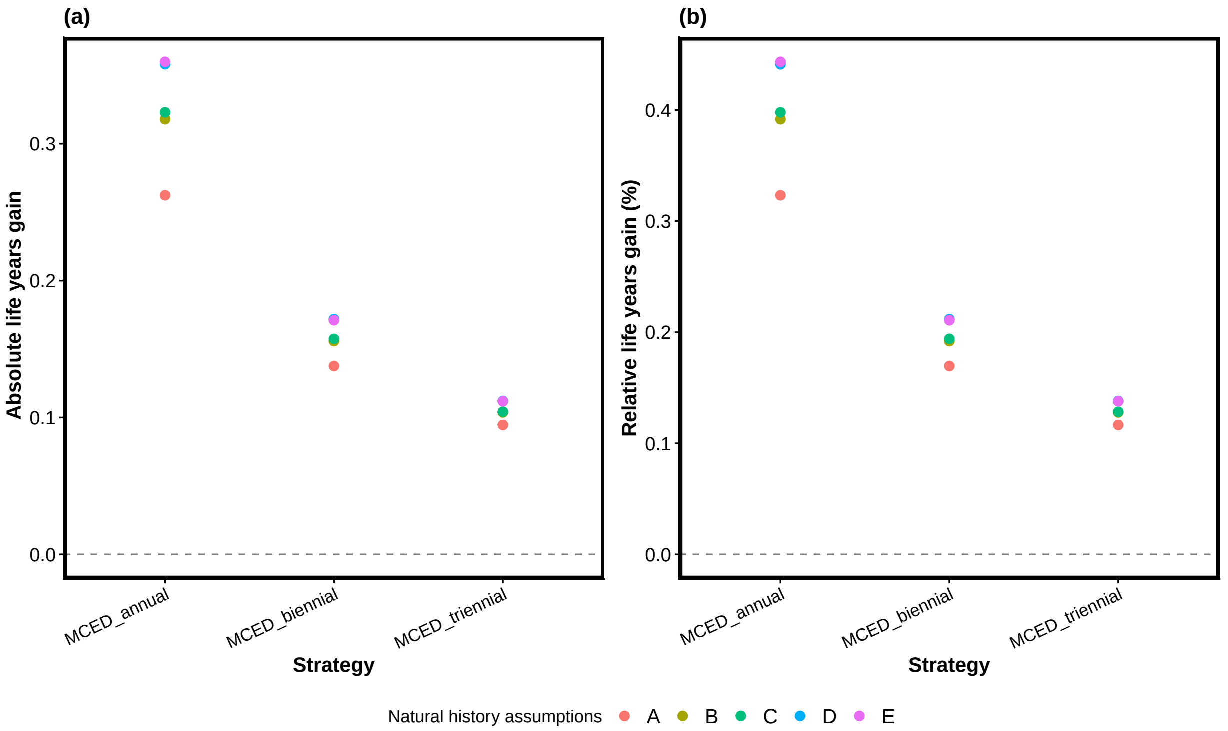

Supplemental Figure 8: Absolute and relative life years gain across screening strategies under optimistic survival assumptions.
